# Failure to balance social contact matrices can bias models of infectious disease transmission

**DOI:** 10.1101/2022.07.28.22278155

**Authors:** Mackenzie A. Hamilton, Jesse Knight, Sharmistha Mishra

## Abstract

Spread of transmissible diseases is dependent on contact patterns in a population (i.e. who contacts whom). Therefore, many epidemic models incorporate contact patterns within a population through contact matrices. Social contact survey data are commonly used to generate contact matrices; however, the resulting matrices are often imbalanced, such that the total number of contacts reported by group A with group B do not match those reported by group B with group A. While the importance of balancing contact matrices has been acknowledged, how these imbalances affect modelled projections (e.g., peak infection incidence, impact of public health measures) has yet to be quantified. Here, we explored how imbalanced contact matrices from age-stratified populations (<15, 15+) may bias transmission dynamics of infectious diseases. First, we compared the basic reproduction number of an infectious disease when using imbalanced versus balanced contact matrices from 177 demographic settings. Then, we constructed a susceptible exposed infected recovered transmission model of SARS-CoV-2 and compared the influence of imbalanced matrices on infection dynamics in three demographic settings. Finally, we compared the impact of age-specific vaccination strategies when modelled with imbalanced versus balanced matrices. Models with imbalanced matrices consistently underestimated the basic reproduction number, had delayed timing of peak infection incidence, and underestimated the magnitude of peak infection incidence. Imbalanced matrices also influenced cumulative infections observed per age group, and the projected impact of age-specific vaccination strategies. For example, when vaccine was prioritized to individuals <15 in a context where individuals 15+ underestimated their contacts with <15, imbalanced models underestimated cumulative infections averted among 15+ by 24.4%. We conclude stratified transmission models that do not consider reciprocity of contacts can generate biased projections of epidemic trajectory and impact of targeted public health interventions. Therefore, modellers should ensure and report on balancing of their contact matrices for stratified transmission models.

**AUTHOR SUMMARY:** Transmissible diseases such as COVID-19 spread according to who contacts whom. Therefore, mathematical transmission models – used to project epidemics of infectious diseases and assess the impact of public health interventions – require estimates of who contacts whom (also referred to as a contact matrix). Contact matrices are commonly generated using contact surveys, but this data is often imbalanced, where the total number of contacts reported by group A with group B does not match those reported by group B with group A. Although these imbalances have been acknowledged as an issue, the influence of imbalanced matrices on modelled projections (e.g. peak incidence, impact of public health interventions) has not been explored. Using a theoretical model of COVID-19 with two age groups (<15 and 15+), we show models with imbalanced matrices had biased epidemic projections. Models with imbalanced matrices underestimated the initial spread of COVID-19 (i.e. the basic reproduction number), had later time to peak COVID-19 incidence and smaller peak COVID-19 incidence. Imbalanced matrices also influenced cumulative infections observed per age group, and the estimated impact of an age-specific vaccination strategy. Given imbalanced contact matrices can reshape transmission dynamics and model projections, modellers should ensure and report on balancing of contact matrices.

## INTRODUCTION

Contact patterns (i.e., who contacts whom) are a fundamental component of infectious disease transmission dynamics. Such patterns, and the role of highly connected subgroups, can determine the size of epidemics, the incidence of infection among subgroups of a population, and whether epidemics emerge and persist [1]. Mathematical models are widely used to study transmission dynamics and evaluate public health interventions; therefore, such models are often structured to consider population and contact heterogeneity [2–6].

Models with population and contact heterogeneity require estimates of contact within and between subgroups, represented through a contact matrix. Data to generate contact matrices are often obtained from contact diaries and surveys, with the POLYMOD social contact study [7] among the most commonly used data sources [2–5]. POLYMOD collected data on daily age-stratified social contacts of almost 8000 individuals across 8 European countries. To enable broader application of POLYMOD data, several techniques were then developed to project contact matrices to other countries [8–10]. For example, Prem et al. [8,9] used a Bayesian hierarchical model with country-specific data on population demographic structure, school enrollment, workforce participation, and household makeup to project POLYMOD contact matrices to 177 different countries around the world.

When using empirical contact data to structure the underlying contact patterns in a population, analysts must consider the balanced (i.e., reciprocal) nature of contacts [11,12]. In reality, the total number of contacts that individuals in subgroup *i* form with individuals in subgroup *j* must be equal to the total number of contacts that individuals in subgroup *j* form with individuals in subgroup *i*, such that:

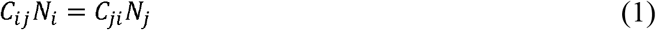

where *C*_*ij*_ is the number of contacts an individual in subgroup *i* forms with individuals in subgroup *j* per day; *N*_*i*_ is the size of subgroup *i;* and likewise for *C*_*ji*_ and *N*_*j*_. However, perfect reciprocity is rarely observed in contact survey data. Imbalances in empirical contact data often arise due to measurement error in survey responses (e.g., recall bias or social desirability bias). However, even when measurement error is absent, imbalances can arise due to selection bias (e.g., differential sampling of subgroups across the network). That is, contact surveys rarely sample from closed, perfectly defined networks, and sampling frames are seldom designed to reflect network structures [11,12].

Numerous mathematical transmission models have used contact data from POLYMOD and Prem et al., but many lack description of methods to handle the imbalanced (i.e., non-reciprocal) nature of these matrices [6,13–15]. As such, it is not always clear if and when age-structured transmission models used balanced or imbalanced contact patterns. Moreover, how imbalanced contact matrices affect modelling projections has yet to be quantified.

We sought to examine how imbalances in contact matrices influence infection transmission dynamics using a theoretical, compartmental transmission model of SARS-CoV-2, stratified by age. Specifically, we examined if and how imbalanced contact matrices influence the estimated: basic reproduction number (*R*_*0*_); temporal epidemic dynamics; cumulative infections among age groups; and impact of age-specific vaccination strategies.

## METHODS

### Study design

We conducted an analytic and simulation (mathematical modeling) study to examine three key characteristics of a model’s underlying transmission dynamics that can be modified by a network structure: the basic reproduction number, the temporal pattern of an epidemic, and the epidemic size. First, we compared the basic reproduction number R_0_ of an epidemic in a population stratified into two age groups (<15 and 15+) when parameterized with imbalanced versus balanced contact matrices across all 177 demographic settings studied by Prem et al. [8]. We used contact matrices from Prem et al. to inform parametrization because of their use in most SARS-CoV-2 transmission models to date.

Next, we conducted a theoretical SARS-CoV-2 simulation study using an SEIR (susceptible-exposed-infectious-recovered) mathematical model in three demographic settings where imbalanced contacts reported by 15+ with <15 were a) larger than (Singapore), b) equal to (Luxembourg), and c) less than (Gambia) balanced contacts between 15+ and <15. We compared the timing and magnitude of peak infection incidence, cumulative infections after one year of seeding, and cumulative infections averted in the context of age-specific vaccination strategies after one year of seeding, when models were parameterized with imbalanced versus balanced matrices.

### Age-stratified social contact data

We obtained age-stratified social contact matrices and population data from Prem et al [8]. Raw matrices were imbalanced, and stratified into 16 age groups, with each matrix element, *C*_*ij*_, representing the mean number of contacts that a person in age group *i* reported with a person in age group *j* per day. To simplify our analysis, we transformed the age-structure of the matrices into two age groups: individuals less than 15 years of age, and 15 and older. Imbalanced contact matrices for these age groups were derived by calculating the population-weighted average contacts per person per day of contributing age groups (e.g. 0-4, 5-9 and 10-14 for the new age group <15) from the raw, imbalanced, contact matrices from Prem et al.

### Derivation of balanced social contact matrices

As has been done previously [10,16,17], we estimated the balanced contacts between individuals in age groups *i* and *j* (*C’*_*ij*_) per day by averaging reported contacts from Prem as follows:

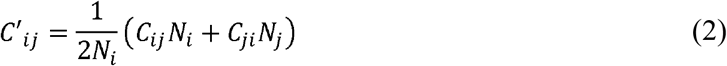

### Derivation of R_0_

We used methodology from Diekmann et al. [18] to calculate R_0_. In brief, R_0_ is the dominant eigenvalue of the next generation matrix (i.e. the number of secondary infectious persons that result in each age group). In a population divided into two age groups, the dominant eigenvalue is the maximum solution of:

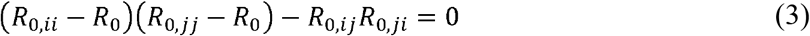

where *i* and *j* denote the two age groups (i.e., <15 and 15+), and *R*_*0,ij*_ is the number of secondary infectious individuals in age group *i* that result from contact with an infectious person in age group *j* in a completely susceptible population, calculated as:

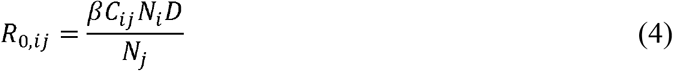

where *β* is the probability of transmission of an infectious disease upon contact, and *D* is the duration of infectiousness. We calculated *R*_*0*_ with imbalanced and balanced matrices from 177 demographic settings studied in Prem [8]. We then calculated a relative R_0_ (*RR*_*0*_) under imbalanced versus balanced conditions where:

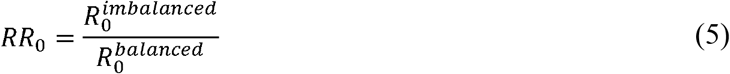

We assumed the probability of transmission and the duration of infectiousness was constant across age groups (and therefore had no impact on the *relative* reproduction number); thus, our estimate of the influence of imbalanced matrices is independent of a specific infectious disease.

### SEIR transmission model

For our simulations, we used a deterministic, compartmental transmission model of SARS-CoV-2 using a simplified SEIR system. Susceptible individuals transitioned to an exposed health state (E) via a force of infection, defined by a probability of contact and probability of transmission per contact with a person in the infectious (I) health state. Individuals in the exposed health state became infectious (I) after a latent period. After an average period of infectiousness, individuals in the infectious health-state moved to the recovered health state (R), where they could not be re-infected. Model equations and details are outlined in **S1 Appendix** material (supplementary equations S1 to S4). **Table 1** summarizes model parameter values.

**Table 1.**
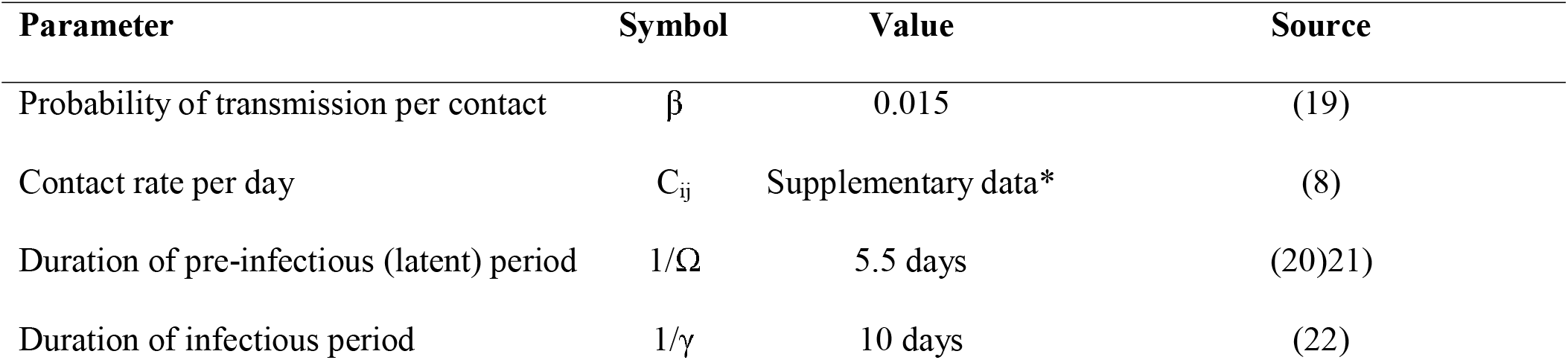
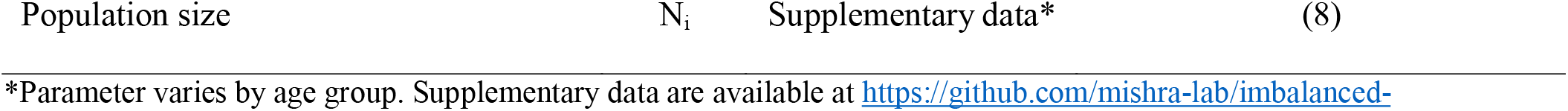
Model parameters.

| Parameter | Symbol | Value | Source |
| --- | --- | --- | --- |
| Probability of transmission per contact | $\beta$ | 0.015 | (19) |
| Contact rate per day | $C_{ij}$ | Supplementary data* | (8) |
| Duration of pre-infectious (latent) period | $1/\Omega$ | 5.5 days | (20)21) |
| Duration of infectious period | $1/\gamma$ | 10 days | (22) |

We made three key assumptions to simplify our model and focus the analysis on the influence of imbalanced matrices. First, we assumed the population size was fixed (i.e. the model simulates a closed system with no births or deaths) to avoid changes in the probability of contact over time. Second, we assumed there were no interventions to mitigate the spread of SARS-CoV-2 (e.g. isolation of infected individuals, reduction in contacts in response to increases in infection rates) as we were interested in isolating the effect of imbalanced matrices rather than infection prevention and control strategies. Finally, we assumed the probability of transmission of, and duration of infectiousness with, SARS-CoV-2 was fixed across age groups to estimate the impact of imbalanced matrices independent of infection properties by age.

### Simulation of SARS-CoV-2 transmission

We simulated SARS-CoV-2 transmission in three demographic settings from Prem et al. [8], where imbalanced contacts that 15+ reported with <15 were: larger than (Singapore), equal to (Luxembourg), and less than (Gambia) balanced contacts between 15+ and <15 (**S1 Fig**). Models were seeded with 1 individual in the infectious state per age group for all simulations. We then compared the magnitude and time to peak incidence, and the percent difference in cumulative infections 1 year after seeding, when models were parameterized with imbalanced versus balanced matrices.

#### Transmission impact of a targeted public health intervention

To explore the influence of imbalanced matrices on the impact of prioritized public health interventions, we simulated two age-specific SARS-CoV-2 vaccination scenarios in all models: one in which vaccines were administered to individuals <15, and another where vaccines were administered to individuals 15 and older. We assumed 50% of the vaccinated age group were immune prior to seeding, and all vaccinated individuals could not be infected (i.e. were permanently immune). We compared cumulative infections overall and per age group in the presence and absence of vaccination over 1 year, to calculate cumulative infections averted from vaccination. Then, we calculated the percent difference in cumulative infections averted between models parameterized with imbalanced versus balanced matrices.

### Validation analyses

To validate robustness of findings, we conducted two additional analyses. First, we assessed how imbalanced matrices affected R_0_ in a population stratified into different age groups: individuals less than 40, and 40 and older. Next, we assessed how imbalanced matrices affected R_0_ when biases in raw contact matrices were opposite to original observations from Prem et al. [8]. For example, if reported contacts between <15 and 15+ were larger than balanced contacts between <15 and 15+ (e.g. Gambia), we forced <15 to underestimate contacts with 15+. We conducted this analysis to assess how systematic bias in contact patterns from Prem may have influenced our results.

## RESULTS

### Imbalance in contact matrices by demographic setting

In comparison to balanced matrices, imbalanced matrices from countries with older populations overestimated total contacts reported by 15+ with <15 (**Fig 1A, red**), and underestimated total contacts reported by <15 with 15+ (**S2A Fig, blue**). The opposite pattern was observed in countries with younger populations, where contacts reported by <15 with 15+ were overestimated (**S2A Fig, red**) and contacts reported by 15+ with <15 were underestimated (**Fig 1A, blue**). For example, in Singapore (median age 42.2 years) the number of imbalanced contacts reported by 15+ with <15 were 1.5 times balanced contacts between 15+ and <15, whereas in Gambia (median age 17.8 years), imbalanced contacts reported by 15+ with <15 were 0.45 times balanced contacts between 15+ and <15.

**Fig 1.**
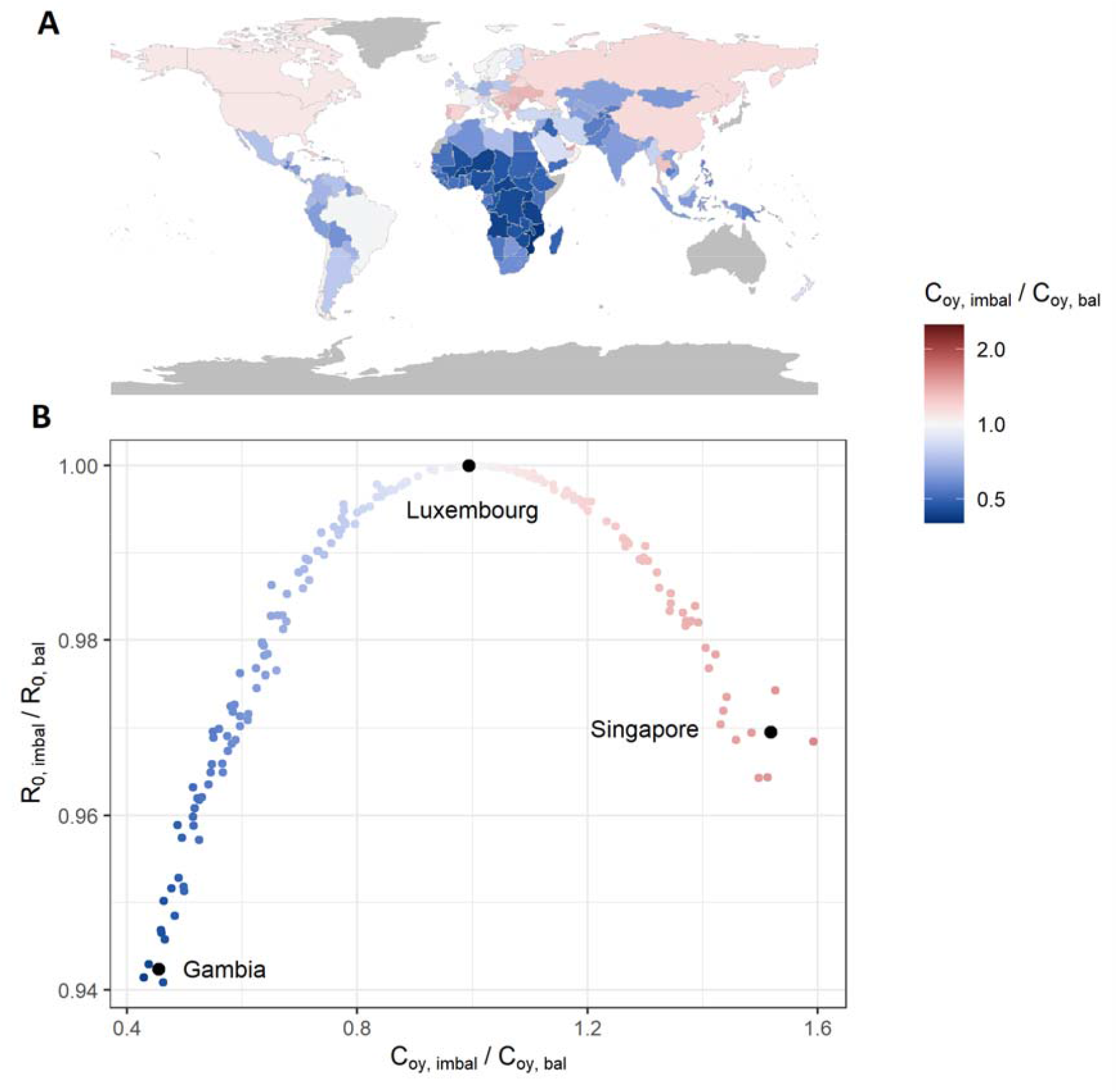
Models with imbalanced contact matrices underestimate R_0_. (A) Direction and magnitude of imbalance in synthetic contact matrix per country. (B) Underestimation of R_0_ in models with imbalanced contact matrices. R_0_, basic reproduction number; C, population contact rate; o, “old”, 15+; y, “young”, <15; imbal, imbalanced; bal, balanced.

### Influence of imbalanced contact matrix on R_0_ and epidemic trajectory

In comparison to models with balanced matrices, models with imbalanced matrices consistently underestimated R_0_ (**Fig 1B, S2B Fig**). For example, R_0_ was 5.7% and 3.1% smaller in Gambia and Singapore respectively, when matrices were imbalanced versus balanced. Models with imbalanced matrices also underestimated the magnitude of, and had delayed time to, peak incidence of SARS-CoV-2 (**Fig 2**). Peak incidence was most dampened and delayed among the age group that underestimated their contacts (i.e., 15+ in Gambia and <15 in Singapore).

**Fig 2.**
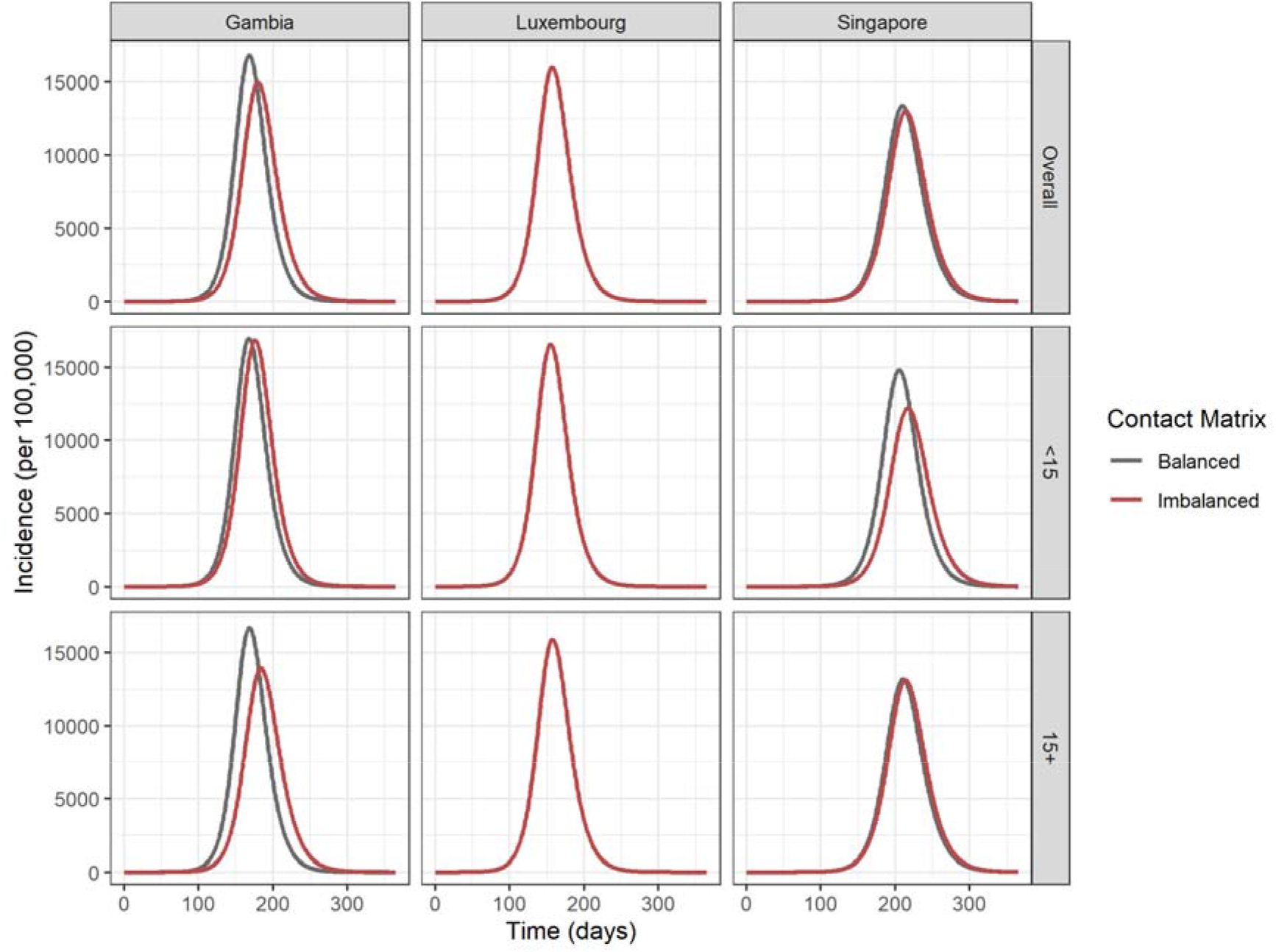
Bias in SARS-CoV-2 epidemic trajectory overall and among age groups according to imbalance in synthetic contact matrix. Models were run among a closed population for 1 year in the absence of public health interventions. Models were seeded with 1 infected individual per age group.

When imbalanced and balanced contacts between 15+ and <15 were similar, there was minimal influence on R_0_ and on the epidemic trajectory of SARS-CoV-2. For example, in Luxembourg imbalanced contacts reported by 15+ with <15 were 0.99 times balanced contacts; therefore, R_0_ was nearly the same under imbalanced and balanced conditions (% difference in R_0_= 0.0003%).

### Influence of imbalanced contact matrix on cumulative infections after 1 year of transmission

Models with imbalanced contacts consistently overestimated cumulative infections in the age group that overestimated their contacts, and underestimated cumulative infections in the age group that underestimated their contacts (**Fig 3**). For example, cumulative infections were 3.2% larger among <15 and 6.7% smaller among 15+ in imbalanced versus balanced models in Gambia; whereas, cumulative infections were 1.6% larger among 15+ and 10.2% smaller among <15 in imbalanced versus balanced models in Singapore.

**Fig 3.**
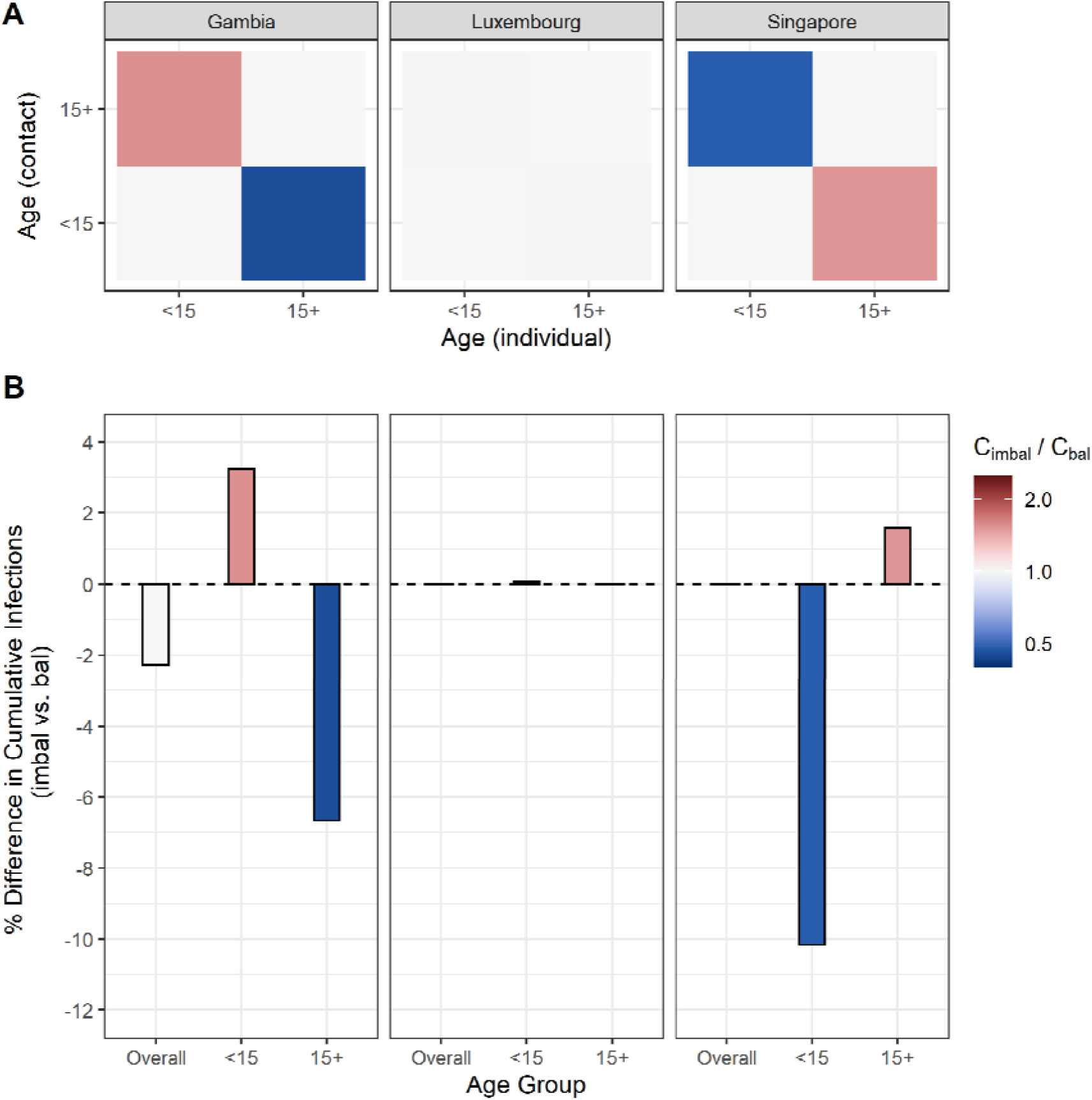
Imbalanced contact matrices bias estimates of cumulative SARS-CoV-2 infections overall and among subgroups. (A) Direction and magnitude of imbalance in synthetic contact matrices from Gambia, Luxembourg and Singapore. (B) Percent difference in cumulative infections overall, and per age group from models parameterized with imbalanced versus balanced contact matrices. One infected individual was seeded per age group per model. Cumulative infections were compared one year after seeding in a completely susceptible and closed population in the absence of public health interventions. Imbal, imbalanced; bal, balanced.

### Influence of imbalanced contact matrix on age-specific vaccination strategies

Imbalanced matrices also directly and indirectly biased projected infections averted from age-specific SARS-CoV-2 vaccination strategies (**Fig 4**). For example, when vaccines were prioritized to individuals <15, imbalanced models underestimated infections averted among 15+ in Gambia (percent difference = -24.4) and overestimated infections averted among 15+ in Singapore (percent difference = 38.8). When vaccines were prioritized to individuals 15+, imbalanced models overestimated infections averted among <15 in Gambia (percent difference = 20.2) *and* Singapore (percent difference = 25.5).

**Fig 4.**
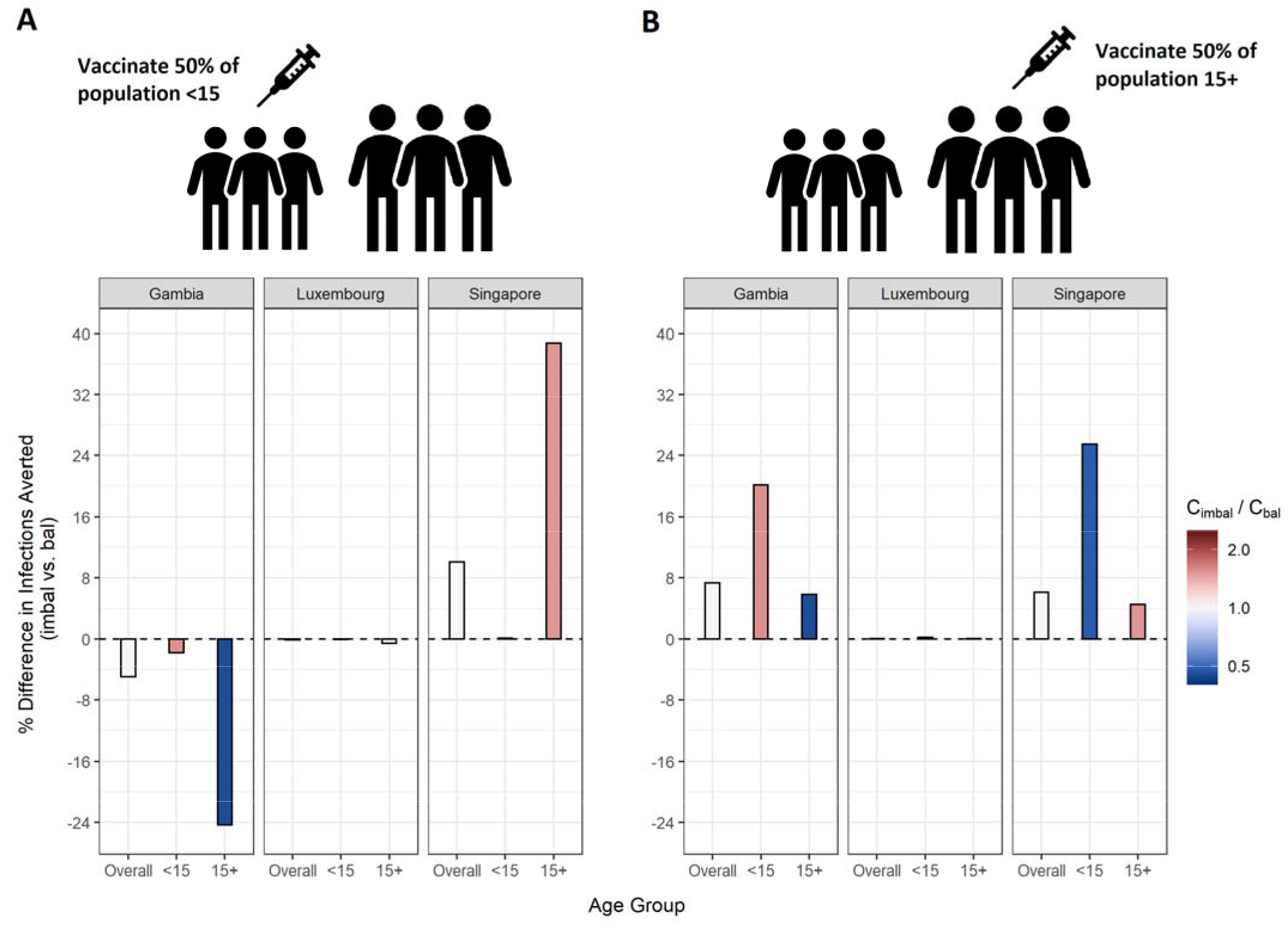
Imbalanced contact matrices bias impact of age-specific SARS-CoV-2 vaccination strategies. (A) Percent difference in cumulative infections averted in models parameterized with imbalanced versus balanced contact matrices when 50% of population <15 was vaccinated (B) Percent difference in cumulative infections averted in models parameterized with imbalanced versus balanced contact matrices when 50% of population 15+ was vaccinated. One infected individual was seeded per age group, per model. Cumulative infections averted were compared one year after seeding in a completely susceptible and closed population, in the absence of additional public health interventions other than vaccination. Imbal, imbalanced; bal, balanced.

### Validation analyses

Our results were robust to changes in stratification of age groups (**S3A Fig**). For example, imbalanced contacts reported by 40+ with <40 were 1.4 and 0.38 times balanced contacts reported by 40+ with <40 in Singapore and Gambia respectively. In these two settings, models with imbalanced matrices underestimated R_0_ by 2.5% and 5.4% respectively.

Results were also robust to assumptions regarding which age group over- or underestimated their contacts (**S3B Fig**). For example, when we forced imbalanced contacts reported by 15+ with <15 to be 0.48 and 1.6 times balanced contacts between 15+ and <15 in Singapore and Gambia respectively (i.e. opposite the original imbalance direction observed in Prem et al.), models with imbalanced matrices still underestimated R_0_ by 3.0% and 5.8% respectively.

## DISCUSSION

Using a combination of analytic and simulation methods, we found that the use of imbalanced contact matrices reshaped the underlying transmission dynamics of SARS-CoV-2. Models with imbalanced matrices consistently underestimated R_0_, leading to: 1) biased time to, and magnitude of peak infection incidence, 2) biased estimates of subgroup specific cumulative infections, and 3) biased impact of age-specific SARS-CoV-2 vaccination strategies. Biases resulting from imbalanced matrices persisted as we varied age group definitions, and as we transformed assumptions regarding which age group over- or underestimated their contacts per demographic setting.

The finding that R_0_ is always smaller when models are parameterized with imbalanced versus balanced matrices can be explained mathematically. In simplifying equation 3, we see that R_0_ is monotonically related to the product of R_0,ij_ and R_0,ji_ (proof provided in **S1 Appendix;** supplementary equations S5 to S8). We can also see that R_0,ij_ and R_0,ji_ are proportionate to C_ij_N_i_ and C_ji_N_j_, respectively (i.e., equation 4). Following the isoperimetric theorem for rectangles, given a fixed sum of population contacts between age groups *i* and *j* (i.e., C_ij_N_i_ + C_ji_N_j_), the product of C_ij_N_i_ and C_ji_N_j_ will be maximized when C_ij_N_i_ = C_ji_N_j_ (i.e., equation 1; conditions for balanced mixing). Since we assumed all other parameters in equation 4 were fixed across age groups, and the sum of population contacts was constant between imbalanced and balanced matrices (i.e., equation 2), the product of R_0,ij,_ and R_0,ji_ will maximize when C_ij_N_i_ and C_ji_N_j_ are equal. Therefore, under our model and assumptions, R_0_ will always be largest under balanced conditions.

It may also be intuitive that biases in cumulative infections per age group are related to biases in contact patterns from imbalanced matrices. The number of infections among subgroup *i* is dependent on the “force of infection” (*λ*_*i*_):

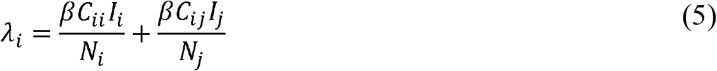

As C_ij_ increases or decreases, the force of infection among subgroup *i* will also increase or decrease, as will the number of infections observed within the subgroup.

Given infection transmission dynamics were biased by imbalanced contact patterns, it was expected that they would also bias impact of subgroup specific public health interventions. This is because biases in contact patterns influence both risk of infection acquisition *and* transmission potential once infected. That is, if a model underestimates contacts that subgroup *i* makes with subgroup *j*, the model also underestimates the transmission potential of subgroup *j* to subgroup *i*. This was most notable when vaccine was administered to 50% of the population <15, where models that underestimated transmission potential of <15 underestimated infections averted among 15+ (i.e. Gambia) and models that overestimated transmission potential of <15 overestimated infections averted among 15+ (i.e. Singapore). Counterintuitively, when vaccine was administered to 50% of the population 15+, imbalanced models overestimated infections averted among <15 in both Gambia and Singapore (i.e. regardless of direction of bias in transmission potential of 15+). We hypothesize imbalanced models from Singapore overestimated infections averted among <15 (despite underestimating transmission potential of 15+) because of indirect bias in infection transmission dynamics. In addition to underestimating transmission potential of 15+, imbalanced models from Singapore overestimated transmission potential of <15. Therefore, in the absence of vaccination, there were more infections observed among 15+ in imbalanced models from Singapore. This provided more opportunity for vaccination to stop transmission of infection from 15+ to <15.

To our knowledge, this is the first study to quantitatively assess bias associated with *imbalanced* contact matrices on compartmental models of infectious diseases. Our work builds on a previous study by Arregui et al. that demonstrated the way in which contact matrices are balanced and projected to new demographic settings can influence the epidemic trajectory observed [10]. The issue of non-reciprocity has been well-recognized in survey data on sexual partnerships, where various methods have been developed to balance sexual partnerships, and balancing is an established component of the modeling of sexually transmitted infections [11,23,24]. However, the importance of balanced contacts has been less discussed, nor established, as part of standard practice and reporting of transmission modeling studies with non-sexually transmitted infections [25]. Given imbalanced matrices can create error in model projections, and models with population heterogeneity are increasingly used to inform public health decisions [26–28], modellers should ensure and report on balancing of their contact matrices.

### Limitations

The simplicity of our model allowed us to quantitatively assess and interpret the impact of imbalanced versus balanced matrices irrespective of other infection transmission parameters. However, our results may vary when studied in open populations (with births, deaths and/or movement of individuals) or when considering infection prevention and control measures such as school closures or isolation procedures. Bias may also vary when considering heterogeneity in biological characteristics, such as immunity to infection, duration of infectiousness, and probability of transmission once infected. For example, we assumed the probability of transmission and duration of infectiousness was constant across age groups. If older individuals were more likely to transmit SARS-CoV-2 than younger age groups, and had a longer duration of infectiousness, they would have greater transmission potential and we may see even greater bias in models that overestimate contacts that <15 made with 15+.

Differences in epidemic dynamics between models with imbalanced versus balanced matrices are also inherently dependent on the method used to balance a contact matrix. We used a population weighted average of reported contacts given the matrices were synthetic [8]. However, when using survey data [e.g., POLYMOD [7]], it is more common to calculate respondent weighted averages of population contacts [10], or use statistical techniques to infer patterns across the population according to participant demographic information [12]. This may change the extent to which raw contacts are considered imbalanced, and thus the magnitude and direction of bias in SARS-CoV-2 R_0_, epidemic trajectory, cumulative infections per subgroup, and impact of prioritized public health interventions.

## Conclusions

We show that compartmental models of infectious diseases parameterized with imbalanced contact matrices may produce biased estimates of initial epidemic characteristics (e.g., R_0_), epidemic trajectory (e.g., timing and magnitude of peak infection incidence), cumulative impact on populations (e.g., cumulative infections per age group), and impact of prioritized public health interventions. To avoid biases in projections, stemming from how the model is parameterized, modellers should account for and report reciprocity of contact matrices in their stratified transmission models.

## Data Availability

Age-stratified contact matrices analyzed in this study were obtained from Prem et. al. Their contact matrices are available for download at https://github.com/kieshaprem/synthetic-contact-matrices. The codes to transform and balance source contact matrices, and generate results for this analysis are available on GitHub at https://github.com/mishra-lab/imbalanced-contact-matrices.

## ACKNOWLEDGMENTS

JK is supported by a doctoral award from the National Sciences and Engineering Research Council of Canada (NSERC CGS-D). SM is supported by a Tier 2 Canada Research Chair in Mathematical Modeling and Program Science (CRC grant number 950-232643). We would like to thank Dr. Korryn Bodner, Linwei Wang, and Ekta Mishra for helpful discussions on biases in mathematical modelling of infection transmission and proofing for a general audience.

