## Appendix for "Failure to balance social contact matrices can bias models of infectious disease transmission"

**SUPPLEMENTARY TEXT**

**SEIR Model Equations**

$$\frac{dS_{i}}{dt}= -\beta S_{i}\sum_{j=1}^{n} (C_{ij}\frac{I_{j}}{N})$$

(S1)

$$\frac{dE_{i}}{dt}= \beta S_{i}\sum_{j=1}^{n} (C_{ij}\frac{I_{j}}{N})- ΩE_{i}$$

(S2)

$$\frac{dI_{i}}{dt}= ΩE_{i}- \gamma I_{i}$$

(S3)

$$\frac{dR_{i}}{dt}= \gamma I_{i}$$

(S4)

**Why is R_0_ maximized when contact matrices are balanced**

We derive basic reproduction numbers (R_0_) in the context of heterogeneous mixing following methodology from Diekmann et al. [1] They prove R_0_ is the dominant eigenvalue of the next generation matrix. Therefore, in a population divided in two age groups, *i* and *j*, R_0_ is the maximum solution of:

$$\left( R_{0ii}- R_{0} \right)\left( R_{0jj}-R_{0} \right)-R_{0ij}R_{0ji}=0$$

(S5)

where R_0ij_ is the number of secondary infectious persons in age group *i* that result from contact with an infectious person in age group *j* in a completely susceptible population. Simplifying equation S5 we obtain:

$$R_{0}^{2}-\left( R_{0ii}+R_{0,jj} \right)R_{0}+\left( R_{0,ii}R_{0,jj}-R_{0,ij}R_{0,ji} \right)=0$$

(S6)

Given equation S6 takes the form ax^2^ + bx + c, we can solve for R_0_ using the quadratic formula:

$$R_{0}= \frac{-b \pm\sqrt{b^{2}-4ac}}{2a} \left\{ \begin{aligned} a=1 \\ b= - R_{0,ii}+R_{0,jj} \\ c=R_{0,ii}R_{0,jj}-R_{0,ij}R_{0,ji} \end{aligned} \right.$$

(S7)

Also recall:

$$R_{0,ij}= \frac{\beta C_{ij}N_{i}D}{N_{j}}$$

(S8)

where $\beta$is the probability of transmission upon contact with a person in an infectious state, D is the duration of infectiousness, N is the population size and C_ij_ is the rate of daily contact an individual in age group *i* has with individuals in age group *j*. Given C_ii_ and C_jj_ do not change when balancing contact matrices, and all other parameters from equation S8 are fixed between imbalanced and balanced conditions, R_0ii_ and R_0jj_ are also fixed. Therefore, the only value that changes in equation S7 from imbalanced to balanced conditions is the latter portion of c: R_0ij_R_0ji_.

Following the isoperimetric theorem for rectangles, given a fixed sum of population contacts between age groups *i* and *j* (i.e., C_ij_N_i_ + C_ji_N_j_), the product of C_ij_N_i_ and C_ji_N_j_ will be maximized when C_ij_N_i_ = C_ji_N_j_ (i.e., balanced conditions). Following equation S8, R_0ij_ and R_0ji_ are directly proportional to C_ij_N_i_ and C_ji_N_j_, and all other parameters are fixed, so R_0ij_R_0ji_ will also maximize when C_ij_N_i_ = C_ji_N_j._ It follows then that c is minimized under balanced conditions and the numerator of equation S7 is maximized under balanced conditions.

**SUPPLEMENTARY FIGURES**

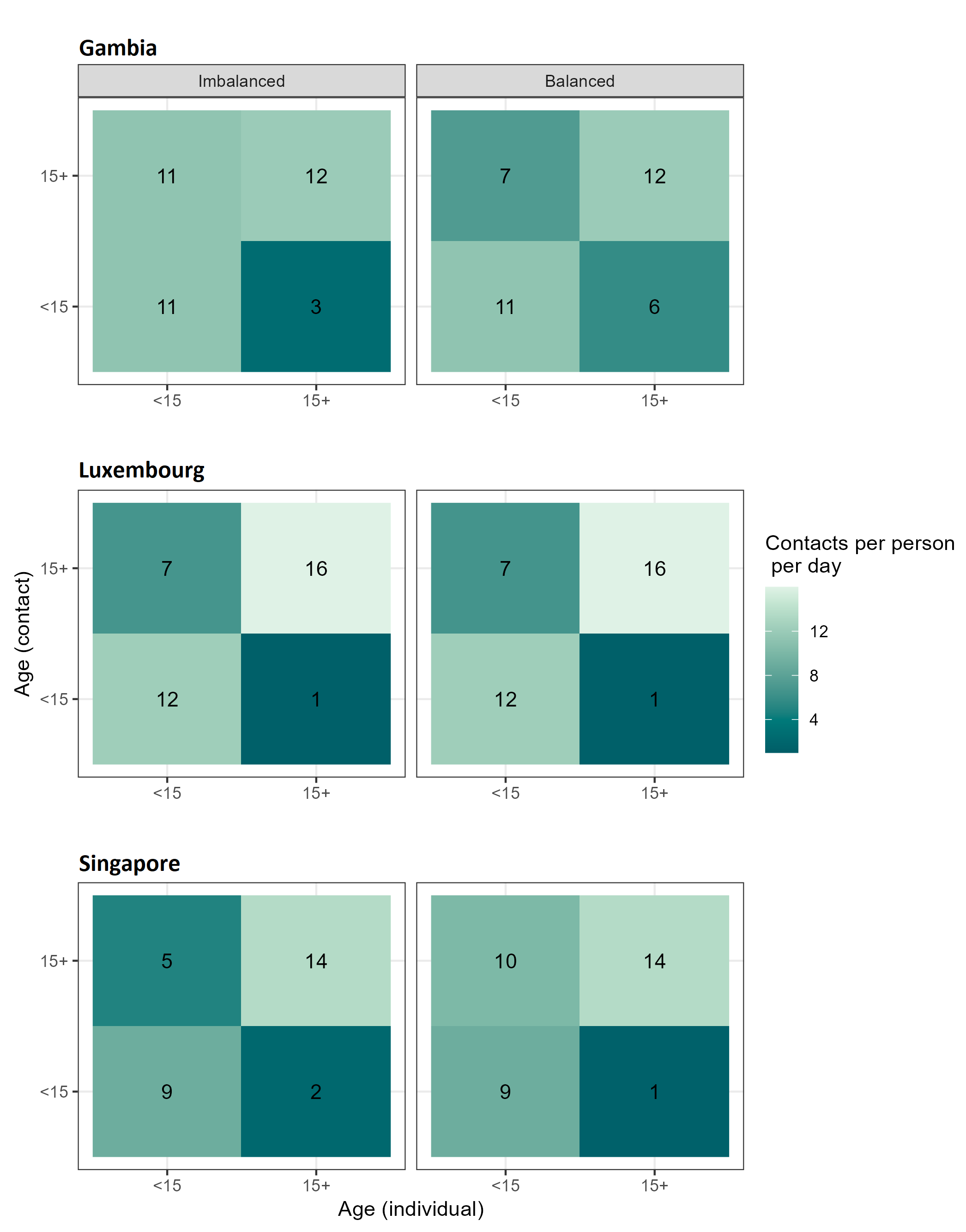

**Fig S1. Imbalanced and balanced age-stratified contact matrices from Gambia, Luxembourg, and Singapore.**

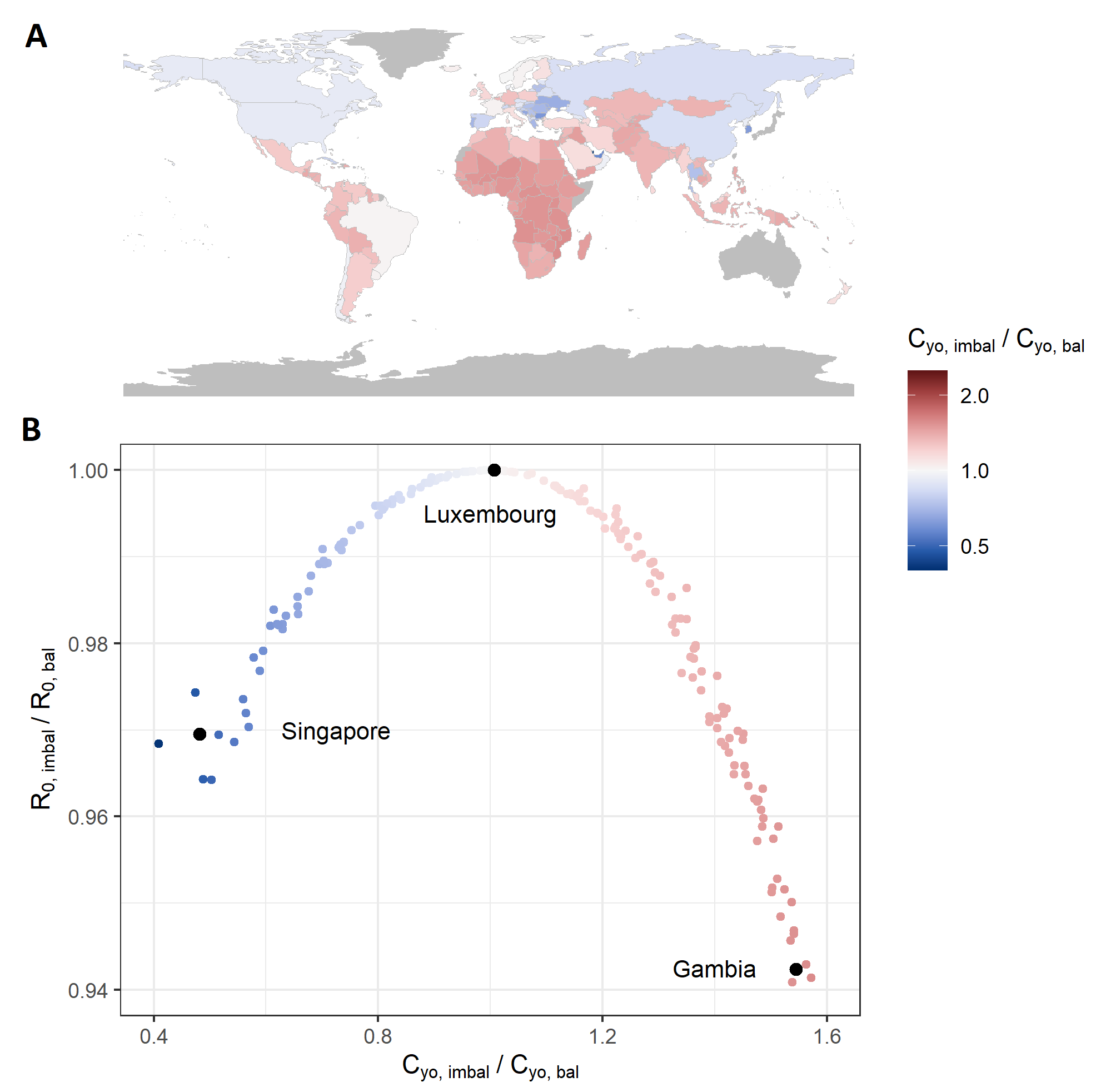

**Fig S2**. **Influence of imbalanced contacts reported by <15 with 15+ on** **R_0_.**

(A) Direction and magnitude of imbalanced in contacts reported by <15 with 15+ per country. (B) Underestimation of R_0_ in models with imbalanced contact matrices. R_0_, basic reproduction number; C, population contact rate; o, 15+; y, <15; imbal, imbalanced; bal, balanced.

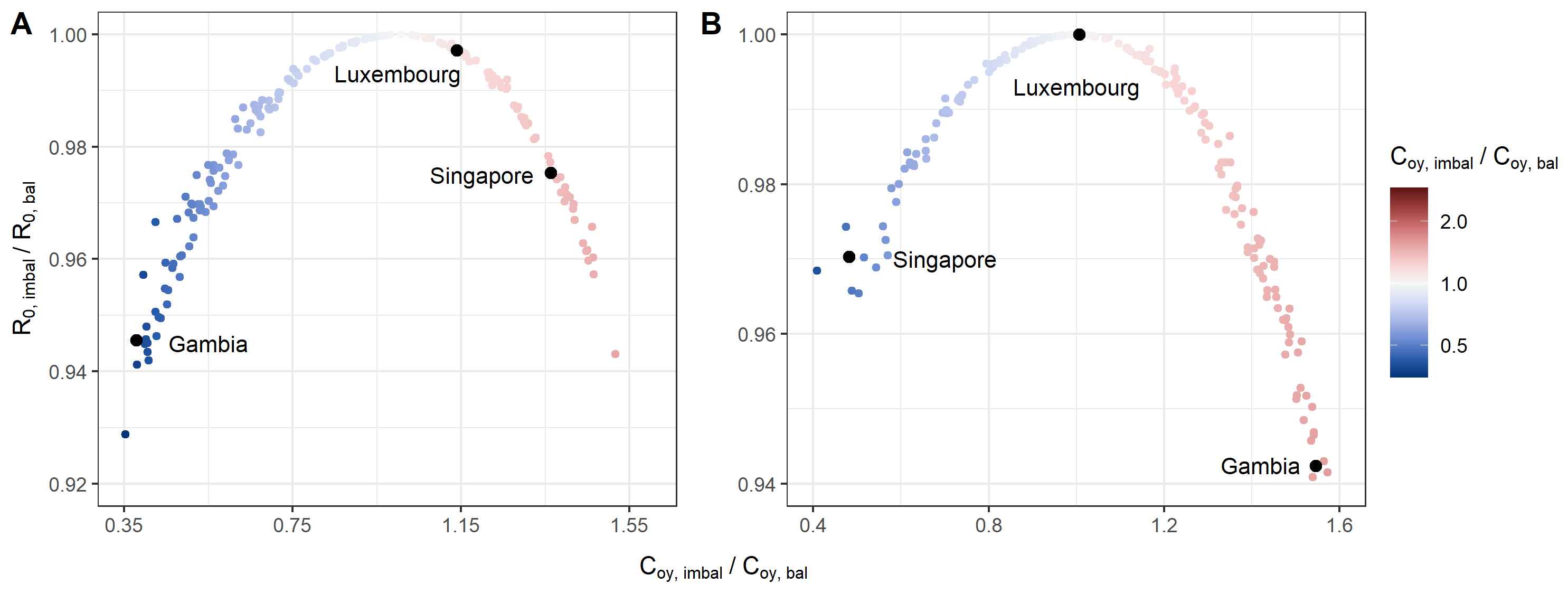

**Fig S3**. **Underestimation of R_0_ is robust to changes in age group stratification and direction of imbalance in synthetic contact matrices.** (A) Bias in R_0_ when using imbalanced synthetic contact matrices from Prem et al. in a population stratified into two age groups: <40 and 40+ years. (B) Bias in R_0_ when using imbalanced synthetic contact matrices with opposite directionality of imbalance in comparison to Prem et al. in a population stratified into two groups: <15 and 15+ years. R_0_, basic reproduction number; C, population contact rate; o, 15+; y, <15; imbal, imbalanced; bal, balanced.
